# Real-time Alerting System for COVID-19 Using Wearable Data

**DOI:** 10.1101/2021.06.13.21258795

**Authors:** Arash Alavi, Gireesh K Bogu, Meng Wang, Ekanath Srihari Rangan, Andrew W Brooks, Qiwen Wang, Emily Higgs, Alessandra Celli, Tejaswini Mishra, Ahmed A. Metwally, Kexin Cha, Peter Knowles, Amir A Alavi, Rajat Bhasin, Shrinivas Panchamukhi, Diego Celis, Tagore Aditya, Alexander Honkala, Benjamin Rolnik, Erika Hunting, Orit Dagan-Rosenfeld, Arshdeep Chauhan, Jessi W Li, Xiao Li, Amir Bahmani, Michael P Snyder

## Abstract

Early detection of infectious disease is crucial for reducing transmission and facilitating early intervention. We built a real-time smartwatch-based alerting system for the detection of aberrant physiological and activity signals (e.g. resting heart rate, steps) associated with early infection onset at the individual level. Upon applying this system to a cohort of 3,246 participants, we found that alerts were generated for pre-symptomatic and asymptomatic COVID-19 infections in 78% of cases, and pre-symptomatic signals were observed a median of three days prior to symptom onset. Furthermore, by examining over 100,000 survey annotations, we found that other respiratory infections as well as events not associated with COVID-19 (e.g. stress, alcohol consumption, travel) could trigger alerts, albeit at a lower mean period (1.9 days) than those observed in the COVID-19 cases (4.3 days). Thus this system has potential both for advanced warning of COVID-19 as well as a general system for measuring health via detection of physiological shifts from personal baselines. The system is open-source and scalable to millions of users, offering a personal health monitoring system that can operate in real time on a global scale.

## Introduction

Early detection of infectious disease enables timely intervention, both to stop transmission and address symptoms. Traditionally, detection has been limited to symptom onset when larger physiological disturbances warrant medical attention. For respiratory viral infections, this is typically several days to over one week post-infection while asymptomatic infections are likely to not be detected at all ^1-3^. When symptom onset does occur, it is usually followed by either an oral or skin temperature measurement or is more definitively diagnosed using a biochemical test such as antigen detection or PCR ^4,5^.

Wearable devices such as smartwatches have the potential to monitor individuals continuously in real time and thus provide early detection of respiratory illnesses and other infections ^6-11^. These devices can collect different types of physiological data such as heart rate, step counts, sleep, and temperature. Recent studies have shown that wearables can be used to identify early signs of infectious diseases such as Lyme disease ^6^ or respiratory viral infections, including COVID-19 ^7-10^, and may even permit pre-symptomatic detection ^6,7^. These respiratory viral infection studies have focused primarily on detection at symptom onset and, even in the case of pre-symptomatic detection, were performed retrospectively. Whether respiratory viral infections and other stress events can be prospectively detected has not been examined nor has a system been developed for performing this at scale. An early detection approach using a monitoring and alerting system would enable early self-isolation, treatment, and allocation of healthcare resources and as such could be an invaluable tool for containing pandemics.

In this study we created the first large-scale real-time monitoring and alerting system for detecting abnormal physiological events, including COVID-19 infection onset, using agnostic algorithms across different types of smartwatches. We designed a novel algorithm capable of detecting outlier measurements associated with physiological stresses in real time, including COVID-19 and other respiratory illnesses, and generating alerts for the device wearer. For pre-symptomatic cases, we show that the system identifies 77% of COVID-19 illness at or before the onset of symptoms. It also identifies asymptomatic cases and signals resulting from vaccination. An association between symptoms and signals was investigated.

## Results

### Study Overview

We constructed a highly secure, real-time alerting system for detecting abnormal periods of stress such as viral infections using wearable devices (Fig. 1A) and conducted a test study approved by the Stanford University Institutional Review Board (protocol number 57022). The system involves participant enrollment through a secure REDCap ^12^ e-consent system as a plugin to the study app, MyPHD ^13,14^. After connecting the smartwatch through the app, wearable data (heart rate, step count, and sleep analysis), and health information (e.g., surveys of illness, symptom, medication, vaccination) were collected and securely transferred in real time to the cloud for further analysis. Three online infection detection algorithms (NightSignal, RHRAD, and CuSum) were hosted on the cloud and the results of one of them, NightSignal algorithm were returned back to the participants in the form of real-time alerts (i.e., red or green alert per day; an example of a signal associated with the alerts in a confirmed COVID-19 positive individual is shown in Fig. 1B). Participants were expected to annotate the alerts via various surveys (COVID-19 test, activities, symptoms, etc). Medical recommendations (e.g. self-isolation, get tested) were not allowed under our IRB protocol.

**Figure 1.**
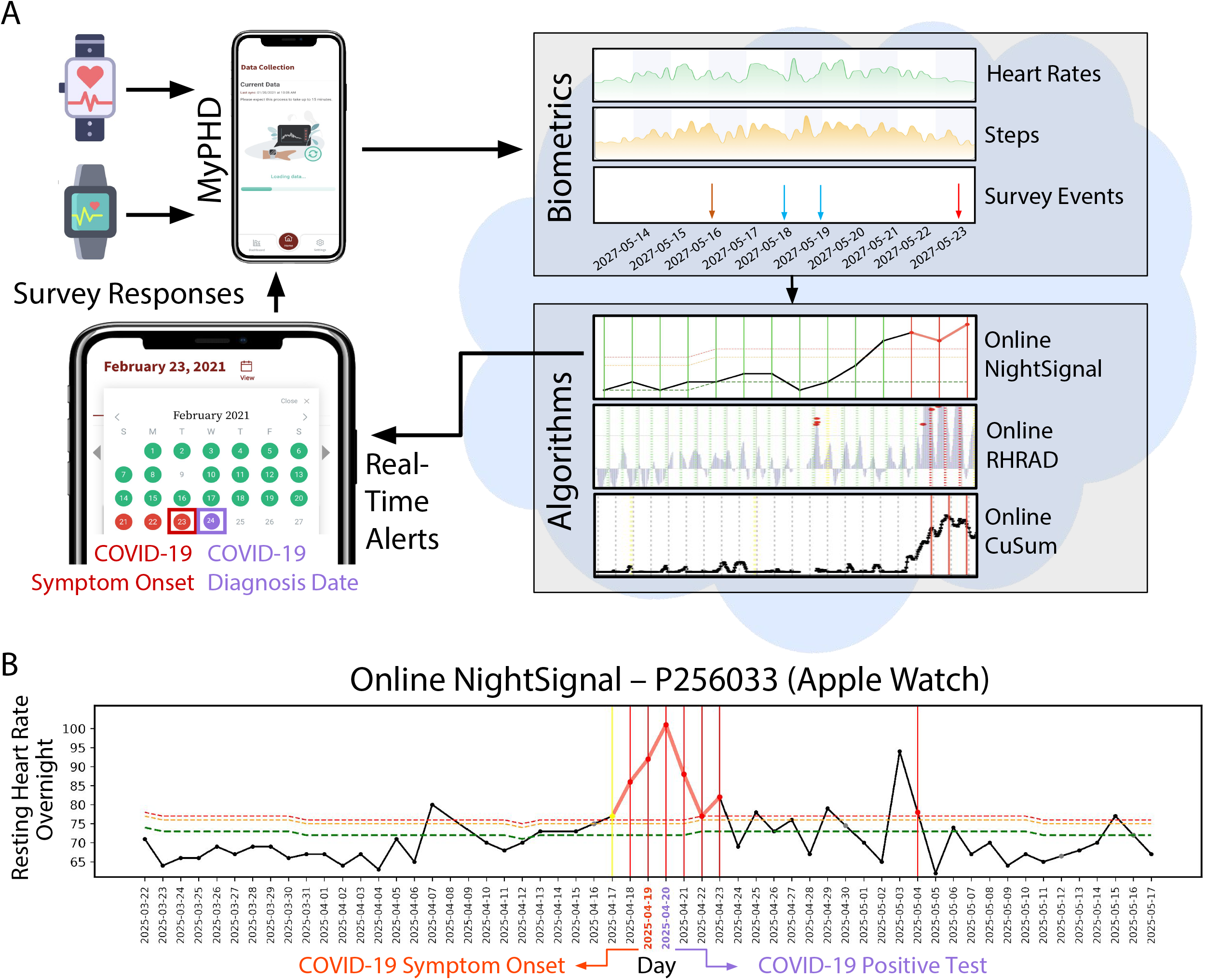
Study overview. **(A)** Participants with a Fitbit and/or Apple Watch were asked to share their wearable and survey data using the study mobile app, MyPHD. The app securely transfers the de-identified data (heart rates, steps, and survey events) to the back-end for real-time analysis. On the back-end, three online infection detection algorithms were deployed and the result from one of the algorithms (online NightSignal) is returned back to the participants in the form of red (indicating abnormal changes in resting heart rate overnight) and green (indicating normal resting heart rate overnight) alerts on the app. **(B)** A real-world example for real-time pre-symptomatic detection of COVID-19 using the online NightSignal algorithm for a participant with Apple Watch. The alerts get triggered two days before the symptom onset date and continue up to three days after the diagnosis date.

We enrolled a total of 3,246 participants between 27 November 2020 and 20 March 2021, of whom 2,112 had wearables data. Of these, 1,016, 950, and 93 wore Fitbit, Apple Watch, or Garmin watches, respectively, and the remaining wore other devices (Supplementary Fig. S1). During the study, 2,076 participants received daily real-time alerts for physiological changes, and 2,075 participants filled in at least one survey in order to annotate the alerts. Among all participants, 207 individuals reported COVID-19 positive test results (67 were confirmed via written documentation of their test result or verbal confirmation). Of those participants reporting COVID-19 positive tests, 68 had sufficient wearable data around the time of COVID-19 infection (35 Fitbit cases and 33 Apple Watch cases - Supplementary Tabble 1). Forty of these cases were retrospective where individuals tested positive prior to enrollment; in 28 cases, the individuals tested positive after enrollment.

We also analyzed wearable data for three other categories of participants: 1) COVID-19 negative individuals: 1,213 participants who reported a COVID-19 negative test and never had a positive test; 2) Potentially healthy individuals: 1,825 participants without any COVID-19 test report; 3) Vaccinated individuals, 189 participants who received the COVID-19 vaccine (Moderna or Pfizer-BioNTech), among them, 182 participants were fully vaccinated (i.e. both doses, Supplementary Tabble 1). Vaccinated individuals could be either COVID-19 positive, negative, or potentially healthy.

Participants who received an alert (or symptoms/activities) were expected to provide a description of their diagnosis, symptoms and activities during that period. Diagnoses included COVID-19, Adenovirus, Influenza, etc. Symptoms included cough, fever, headache, etc, as well as severity (1-mild to 5-very severe). Activities included intense exercise, alcohol, travel, stress, and other lifestyle factors which could alter physiological signals. Overall, 678 of 4,217 alerting periods (two or more consecutive red alert days) were annotated by participants. 51 alerting periods were associated with COVID-19 illness periods (14 days before to 21 days after symptom onset).

### COVID-19 triggers real-time alerts

We used three independent real-time alerting algorithms capable of detection and tracking of physiological changes due to infections such as COVID-19. Two algorithms, online RHRAD and CuSum, extended from our previous work ^7^, detect abnormal deviations from the baseline in resting heart rates from two approaches in anomaly detection (see Methods). These two approaches have been applied on dense Fitbit data, and have the potential to report an alarm at hourly resolution. Although they can detect anomalies in high resolution, they are computationally intensive. In order to be less computationally intensive and potentially scalable to millions of users with various smartwatches, as well as achieving higher sensitivity while keeping the false-positive rate minimal, we developed a novel lightweight algorithm (NightSignal) that uses a deterministic finite state machine (FSM)^15^ based on overnight resting heart rate (RHR) (see Methods and Supplementary Fig. S2). For each individual we use the streaming median of average overnight RHR as the baseline, and raise real-time daily alerts as deviations from baseline as defined by the alerting state machine (see methods). Deviations in successive nights (31 hr) triggers an alert. This algorithm runs on both the Fitbit and Apple Watch and its features are presented in Supplementary Fig. S2. The NightSignal method has the highest sensitivity (78%) compared to CuSum (54%) and RHRAD (44%), but produces the same rate of false (non-COVID-19) signals.

### Real-time pre-symptomatic and asymptomatic detection

We first examined how well the real-time alerting system detects COVID-19 at an early, pre-symptomatic stage and in asymptomatic cases. Fig. 2 shows two examples of prospective detection confirmed by COVID-19 positive tests (a Fitbit case on the top and an Apple Watch case on the bottom) in which we detected elevated NightSignal alerts starting at least 3 and 10 days before symptom onset, respectively. The screenshots of the MyPHD app at the top of Fig. 2 show the real-time alerts that the corresponding participant receives every day. For the Fitbit case, all three algorithms raise alerts before the symptom onset.

**Figure 2.**
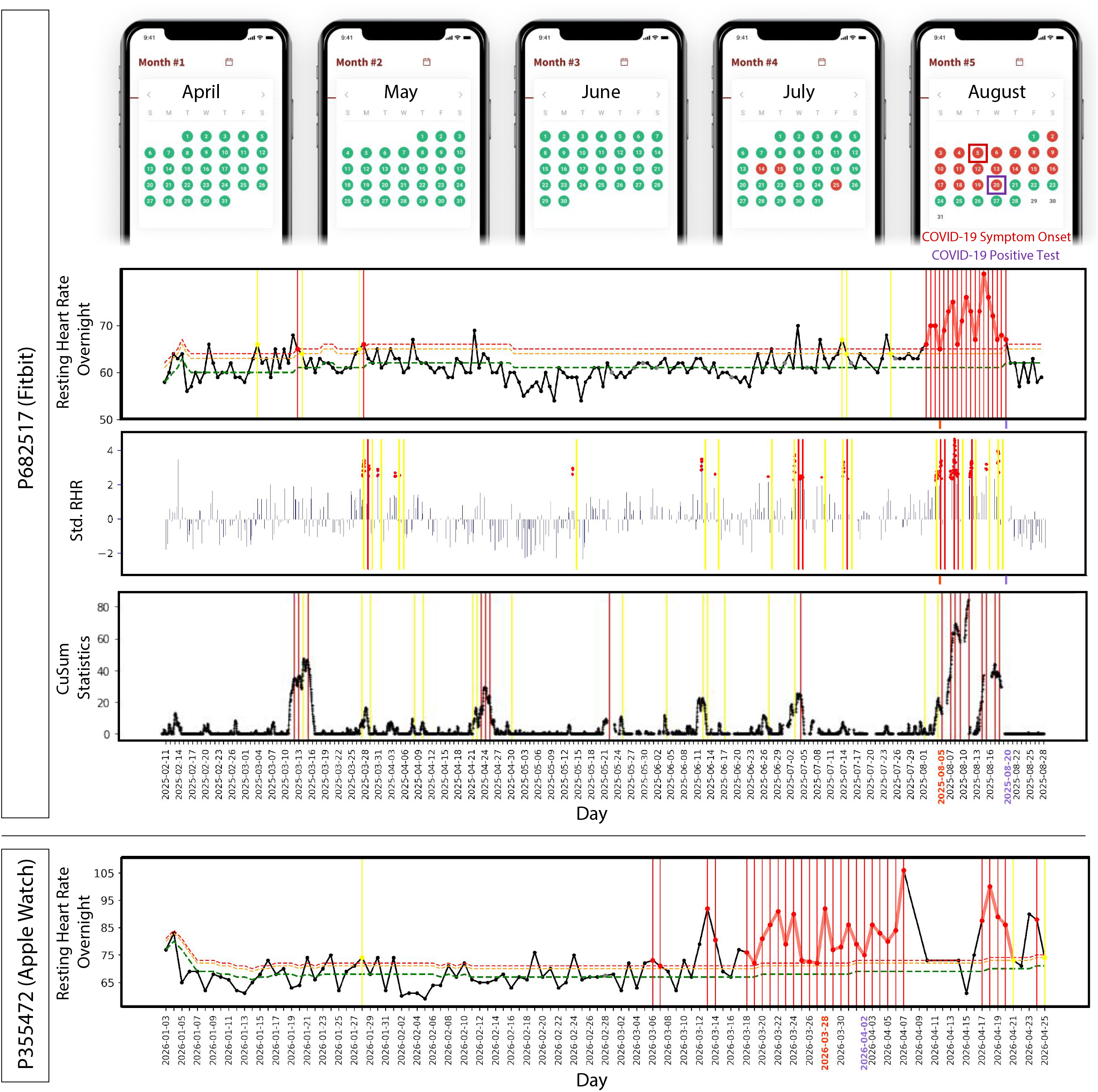
Examples of COVID-19 real-time pre-symptomatic detection. Online pre-symptomatic detection for COVID-19 positive participants with Fitbit (top) using online NightSignal, online RHRAD, and online CuSum algorithms respectively, and Apple Watch (bottom) using online NightSignal algorithm. In the top panel, alerts in the NightSignal algorithm get triggered three days before symptom onset and remain on for the following 15 days. In the bottom panel, signals appear at least 10 days before the symptom onset and continue up to 10 days after that.

We also found 10 cases in which individuals reported testing positive but were asymptomatic; direct communication with the participants confirmed they were asymptomatic. Alert signals were found associated with diagnosed tests in eight of the cases. Two examples of asymptomatic COVID-19 positive participants are shown in Fig. 3 for whom the alerts triggered 19 and 7 days before the COVID-19 diagnosis date, respectively.

**Figure 3.**
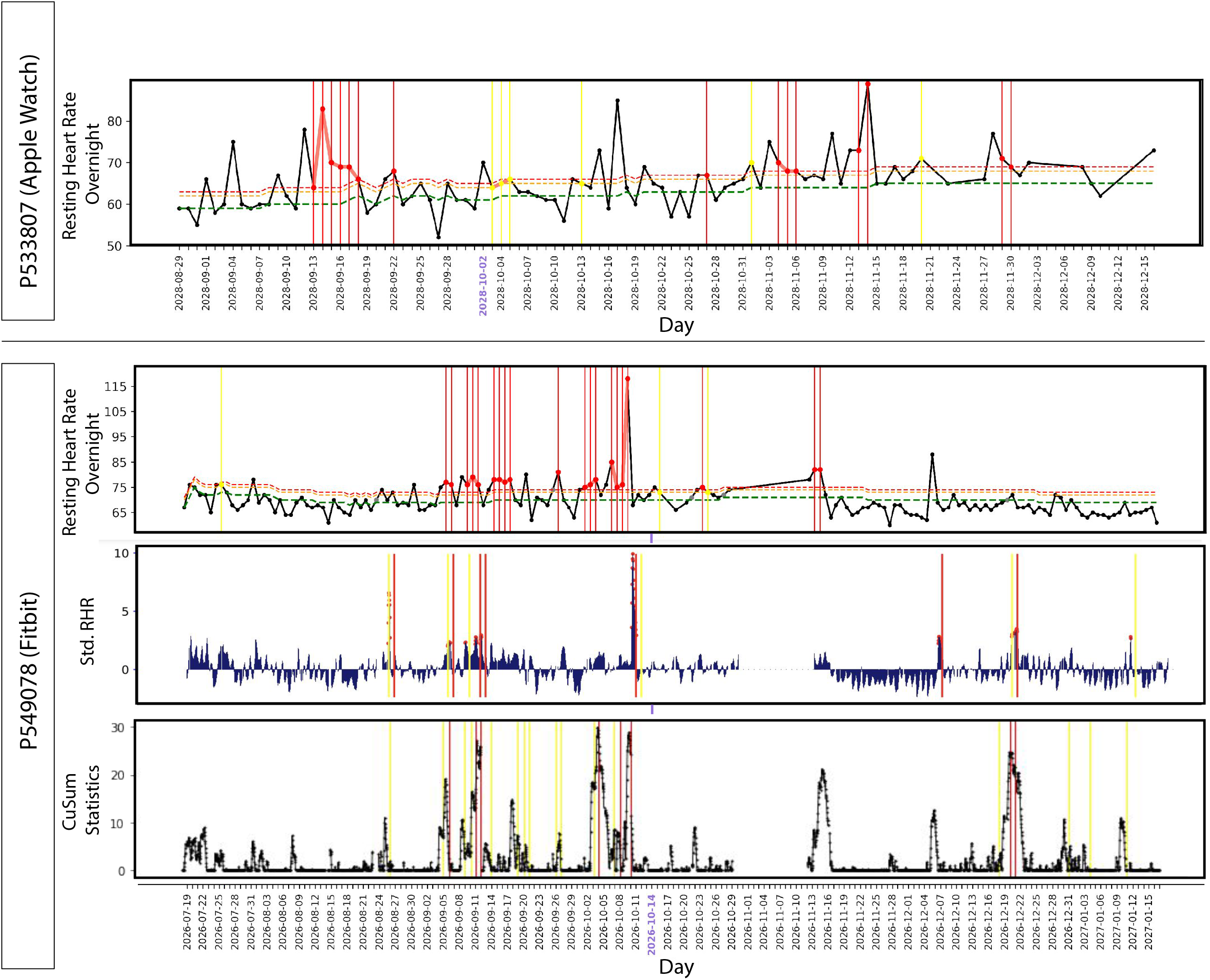
Examples of COVID-19 real-time asymptomatic detection. Online asymptomatic detection for COVID-19 positive participants with Apple Watch (top) using online NightSignal algorithm, and Fitbit (bottom) using online NightSignal, online RHRAD, and online CuSum algorithms respectively.

A summary of alert signals for all pre-symptomatic and asymptomatic cases is shown in Fig. 4. In order to increase detection power, we included the results from 40 retrospective cases where individuals tested positive prior to study enrollment, as well as 28 cases where individuals tested positive after enrollment. Of these, 35 were Fitbit users and 33 were Apple Watch users. For the COVID-19 positive participants, true positives (TP) are defined as the number of cases that received red alerts from the algorithm before or at COVID-19 symptom onset (for pre-symptomatic cases) or diagnosis (for asymptomatic cases) and false negatives (FN) are defined as the number of participants who did not receive any alert within the same infection detection window. Using the COVID-19 negative as well as potentially healthy participants, true negatives (TN) are defined as the number of green alerts that have been correctly sent to these participants and false positives (FP) are the number of red alerts that have been incorrectly sent to these participants during non-COVID-19 periods. Of 68 COVID-19 positive participants (58 symptomatic and 10 asymptomatic), 53 cases received NightSignal alerts at or before symptom onset (for pre-symptomatic cases) or diagnosis (for asymptomatic cases); hence a sensitivity (true-positive rate) of 78% (Fig. 4A and Fig. 4B). The number was similar for Fitbit (74%) and Apple Watch (82%). An additional five cases received alerts within 21 days post-symptom onset, and eight cases did not yield any NightSignal alerts in the infection period (−21 to +21 days around the onset of symptoms). Please note that lack of sufficient wearable data may have been a reason for some of these missed cases.

**Figure 4.**
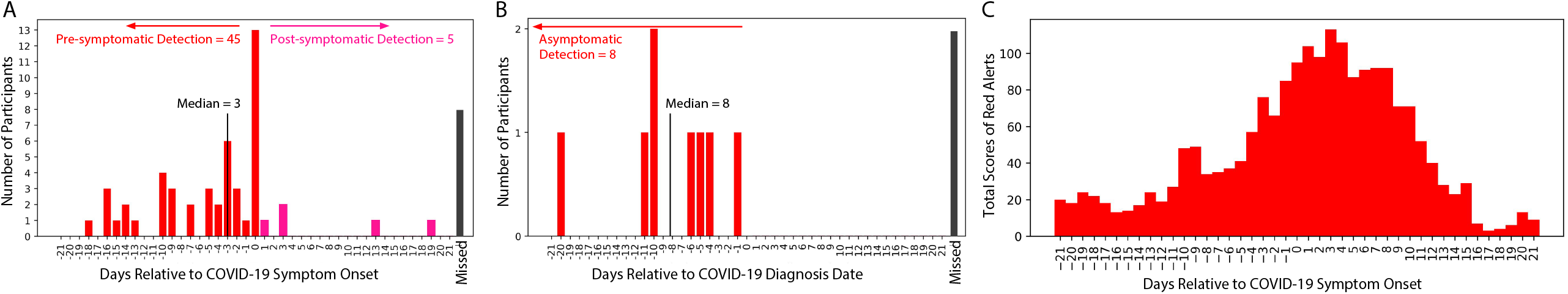
Summary of association of red alerts in the NightSignal algorithm with COVID-19 sick period. **(A)** Association of the initiation of red alerts in the NightSignal algorithm with COVID-19 symptom onset in 58 positive participants with symptoms using a Fitbit or Apple Watch with respect to the detection window of time centered around symptom onset (21 days before to 21 days after symptom onset). NightSignal algorithm achieves pre-symptomatic detection in 45 participants, and post-symptomatic detection in five participants; eight participants did not receive any red alert associated with their COVID-19 symptoms during sickness detection window. **(B)** Association of the initiation of red alerts in the NightSignal algorithm with COVID-19 positive test in asymptomatic participants using a Fitbit or Apple Watch. The plot shows 21 days before and 21 days after the COVID-19 diagnosis date. NightSignal algorithm achieves detection in eight participants; two participants did not receive any red alert associated with their COVID-19 with respect to asymptomatic detection window. **(C)** The plot shows the distribution of scores of red alerts with respect to 21 days before and 21 days after the COVID-19 symptom onset date in positive participants. Red bars indicate the scores of red alerts based on formula (1) below. Let *D = {d*_*1*_, *d*_*2*_, *…, d*_*n*_*}* be a set of days and *R = {d*_*k*_, *d*_*k+1*_, *…, d*_*K+m*_*}* be a set of days where consecutive *k* red alerts have occurred, then the associated red alert score for each day in set R is set to the size of set R (i.e., m+1). Note that the most clustered alerts appear around the symptom onset date. 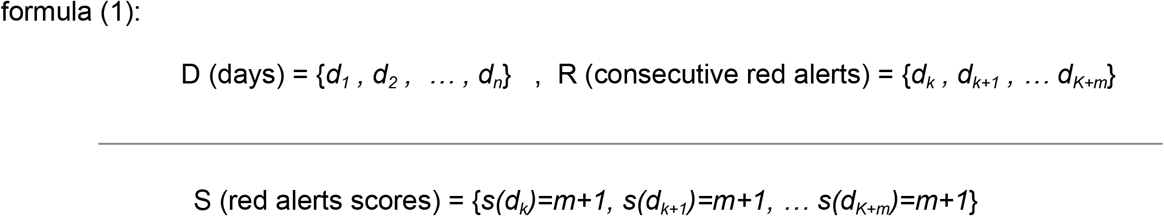

For the case of Fitbit which had hourly data, we were able to analyze the sensitivity of the online RHRAD and CuSum algorithms; these were found to be 12/27 (44%) and 15/28 (54%), respectively. To measure the performance of the algorithms on COVID-19 negative as well as potentially healthy participants, the specificity (true negative rate) of the algorithms defined as TN/TN+FP is as follows: NightSignal 169,980/(169,980+25,610) = 86.9%, Online RHRAD 65,695/(65,695+9,573) = 87.2%, and Online CuSum 85,225/(85,225+12,219) = 87.4%. As described below, these values are with respect to COVID-19 detection--other biologically relevant events could trigger these alerts.

To examine the alerting period relative to symptom onset, we calculated the scores of red alerts based on formula (1) and plotted the distribution with respect to the period of time centered around symptom onset (see Fig. 4C and Methods). Across the 58 symptomatic participants, we observed the maximum number of clustered alerts occur in a window of -4 to +11 days around symptom onset.

### Symptoms and activities raise resting heart rate signals

A wide variety of symptoms are associated with COVID-19 ^16,17^. To examine illness progression, we aggregated symptoms by both severity and number of individuals reporting across 21 days relative to symptom onset (Fig. 5A). Consistent with the literature ^18,19^, most symptoms were evident in the first 7-8 days of illness (fatigue, headache and feeling ill) although fatigue often continued well post symptom onset. An example of an individual with many symptoms, some which persisted for 3 or more weeks, is shown in Fig. 5B.

**Figure 5.**
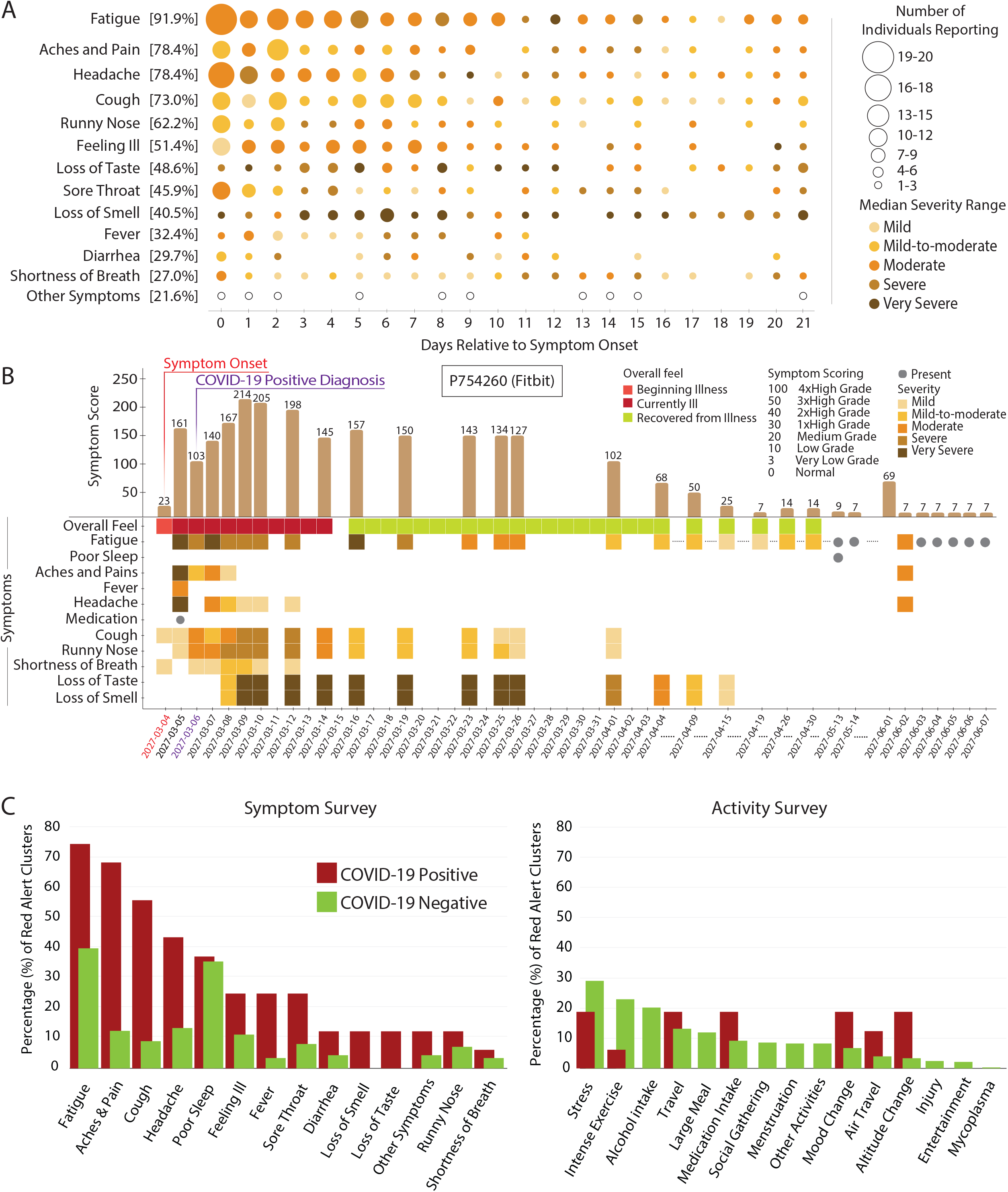
Symptom progression and extent of association with clustered red alerts in COVID-19 sick period. **(A)** Bubble plot representing day by day frequency count of individuals reporting symptoms during the second half of infection detection window (from symptom onset to 21 days later), with the size of the bubble and shading level indicative of the relative magnitude of the frequency count and the median severity respectively. The percentage in the brackets alongside each symptom indicates the aggregate over all the 21 days of the frequency count of individuals reporting that symptom, as a fraction of total symptomatic COVID-19 positive participants (58). Notably, fatigue is the most commonly reported symptom, whereas loss of smell or taste seem to be the highest in terms of severity. **(B)** Illustrative example tracing the symptoms of a COVID-19 positive participant from symptom onset to 21 days after, and continuing on intermittently for additional two months thereafter. For each day, an aggregate symptom score computed as the sum of the relative severities of the symptoms, each weighted by its specificity to COVID-19, shows a bell shaped curve. Also notable is that fatigue stays on as a long hauler symptom. **(C)** The bar plot shows the percentages of red alert periods (from NightSignal algorithm) associated with each symptom/activity as annotated by both COVID-19 positive and negative participants. Except for fatigue and poor sleep, all other symptoms show a wide margin between the higher alert association of the COVID-19 positives and the lower alert association of the COVID-19 negatives.

Out of 4,217 total red alert clusters (at least two or more consecutive red alert days) across the entire study cohort, 678 had annotations by participants. Among them, we examined the red alert clusters annotated by COVID-19 positive participants during the infection detection window (14 days before to 21 days after symptom onset) vis-a-vis those annotated by COVID-19 negative participants (those receiving a negative diagnosis). The distribution of symptoms and activities across the clusters was quite different depending on COVID-19 diagnosis (Fig. 5C). Symptoms such as fatigue and poor sleep were generally present for both COVID-19 positive and negative cases, but aches and pains, headaches, cough and feeling ill were less frequent in the COVID-19 negative cases. Stress, intense exercise, and alcohol intake were activities most commonly associated with red alerts in individuals with COVID-19 negative diagnoses. Furthermore, the mean duration of red alert clusters was higher (4.3 days) for COVID-19 positives than COVID-19 negatives (1.9 days).

Examples of the alerts and symptoms signals are shown longitudinally from a COVID-19 positive case (Fig. 6A), a COVID-19 negative case (Fig. 6B), a diagnosed *Mycoplasma pneumoniae* infection (Fig. 6C), individuals annotating stress or work stress (Fig. 6D-E), repeated alcohol consumption (Fig. 6F), and extended altitude change (Fig. 6G). These results indicate other events can be attributed to red alerts when surveys are regularly reported. It may also be observed that the RHR elevation during the red alert cluster is noticeably higher for the COVID-19 positive case as compared to the COVID-19 negative and Mycoplasma cases. More examples of different categories (COVID-19 positive, negative, and potentially healthy) are shown in Supplementary Fig. S5. Finally, as reported previously, we note that many non-COVID-19 alerts are evident over the winter holidays--more than other times of the year (a 1.4-fold increase in red alerts; Supplementary Fig. S7). In this study, this increase was evident during the pandemic, whereas it was before the pandemic in our previous study, indicating these events occur independent of the pandemic. It is possible that these holiday associated events are due to increased stress, alcohol and/or travel.

**Figure 6.**
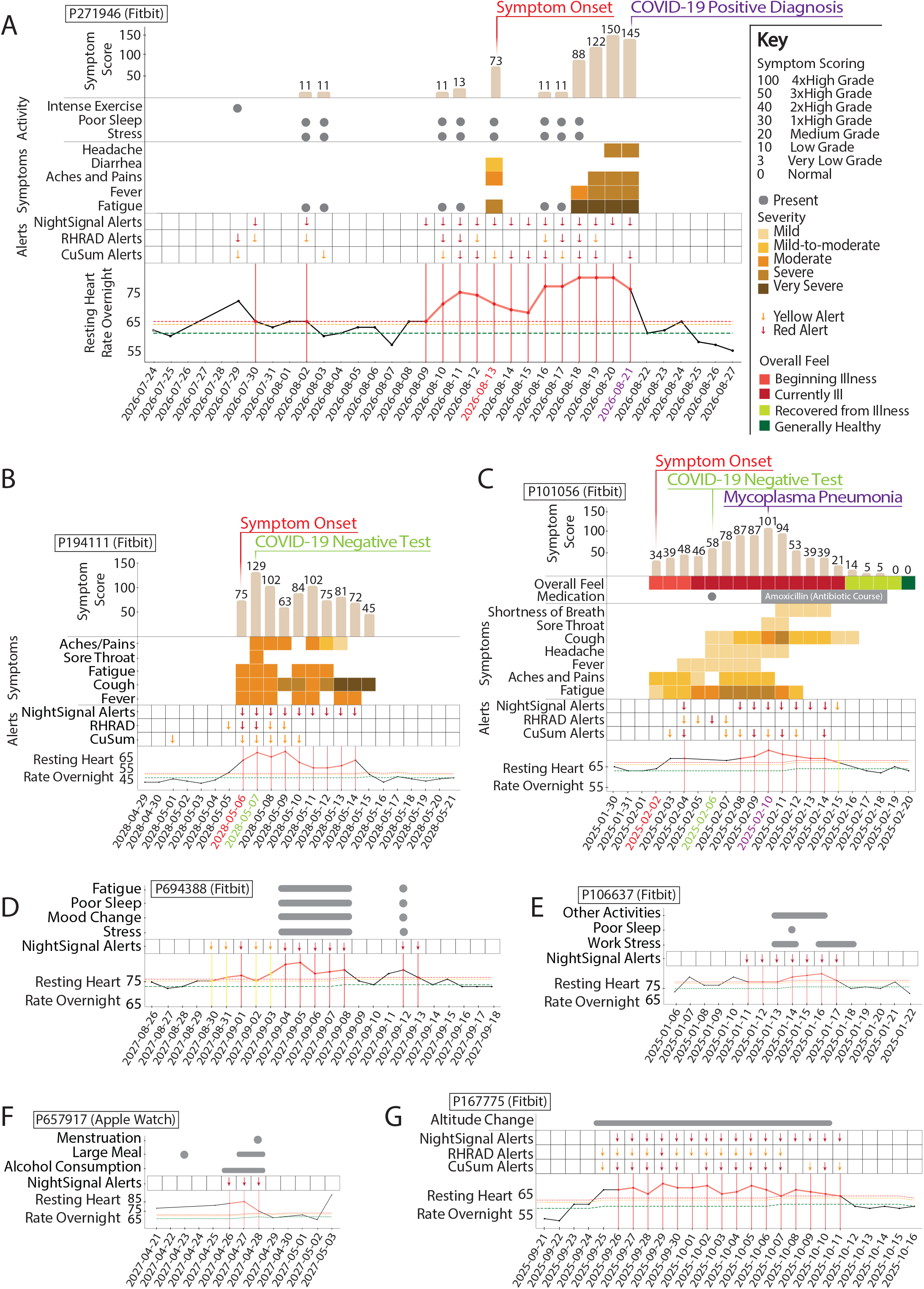
Examples illustrating the association between RHR elevation (as they trigger real time alerts from online NightSignal, online RHRAD, and online CuSum algorithms), and symptoms/activities. **(A)** COVID-19 positive case: Alerts begin prior to symptom onset and continue until diagnosis date. Higher RHR elevations seem to be triggered by severe fatigue, fever and headache. **(B)** COVID-19 Negative case: Alerts even though present throughout the symptom period, the magnitude of RHR elevation is noticeably lower. **(C)** Mycoplasma Pneumonia case: Alerts begin post symptom onset and continue until a day before recovery. Symptom scores follow a neat bell shaped curve. An interesting observation: On the 4th day, the participant got tested for COVID-19 but was negative. However, symptoms continued to increase and when tested on the 8th day for other respiratory infections, Mycoplasma Pneumoniac was detected. On the 14th day (5 days after antibiotic therapy), both symptoms and alerts receded. **(D) and (E)** Stress, poor sleep and mood change correlate well with occurrence of alerts in the COVID-19 Negative individuals. **(F)** Association between repeated alcohol consumption and alerts. **(G)** Association between extended altitude change and alerts.

### COVID-19 vaccination often yields real-time alerts

On December 11, 2020 and December 18, 2020, the U.S. Food and Drug Administration (FDA) issued the first emergency use authorization (EUA) to the Pfizer-BioNTech and Moderna COVID-19 vaccines, respectively ^20,21^. These randomized vaccine clinical trials showed 94%–95% efficacy in preventing COVID-19 illness. Overall, localized side effects (e.g. aches, rash) after vaccination have been shown to be mild; however, moderate-to-severe systemic side effects, such as fatigue and headache were reported by some participants ^21^. In early January, we added COVID-19 vaccination surveys to the study app, MyPHD. As expected, alerts were triggered one to a few days post-vaccination in many participants. Three examples of possible effects of vaccination on NightSignal algorithm detection and symptoms are shown in Fig. 7A. As longitudinally demonstrated in these examples, vaccination can trigger the alerts after both doses or only one dose; however in some cases RHR overnight may increase only for a short period (e.g., one night) and hence no alert is raised. To determine the effect of vaccination on RHR overnight, we analyzed the average RHR overnight for five days before and after the vaccination date. Interestingly, we observed that for the first dose the maximum RHR overnight occurs the night of the vaccination in the case of Pfizer-BioNTech vaccine (an increase was not evident for Moderna); for the second dose it occurs the first or second night after the vaccination for the case of Moderna and Pfizer-BioNTech vaccines, respectively (Fig. 7B).

**Figure 7.**
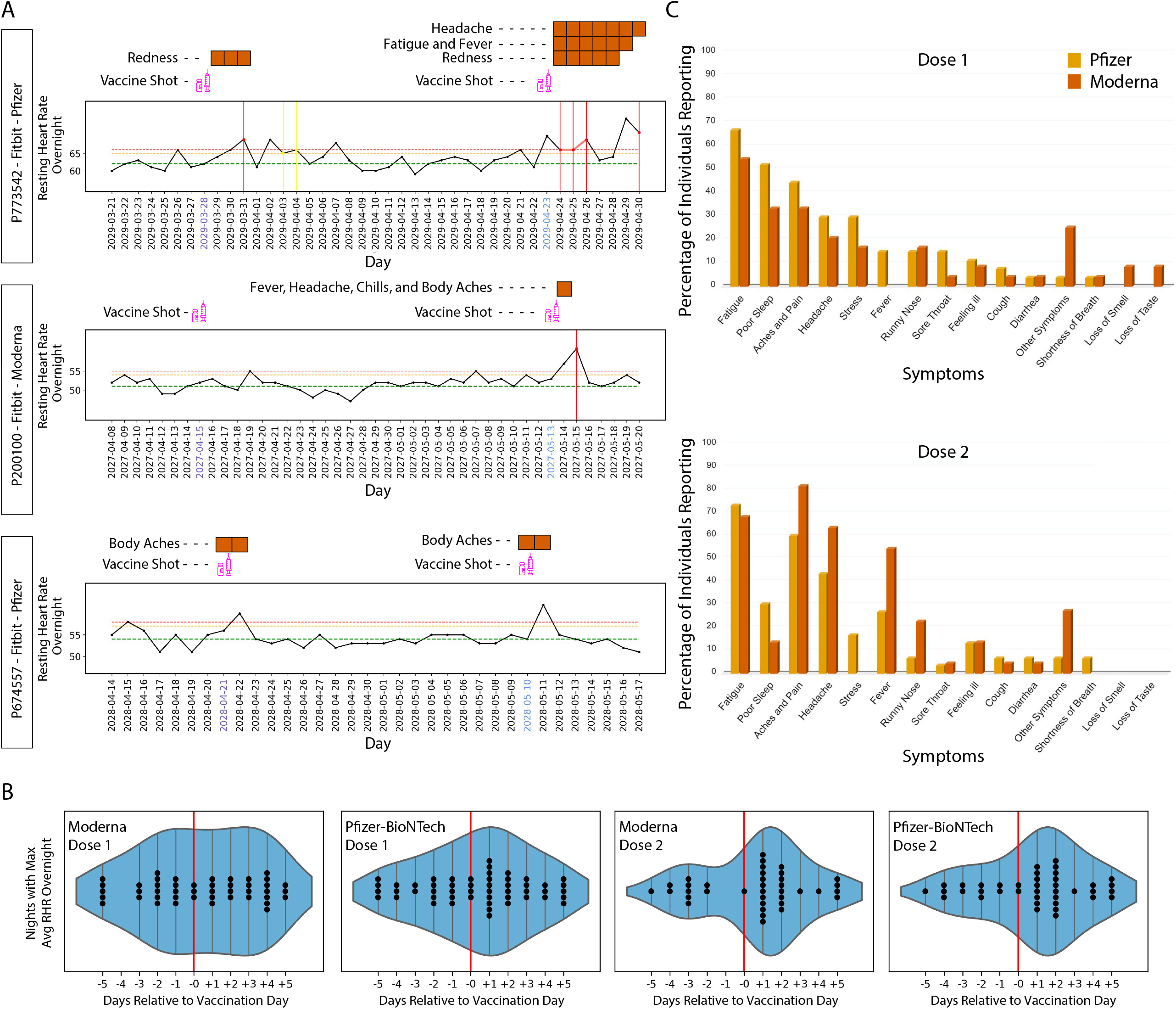
Association of red alerts in the NightSignal algorithm and RHR overnight with COVID-19 vaccination. **(A)** Examples of the effect of COVID-19 vaccination on triggering the alerts in the online NightSignal algorithm. **(B)** The significant effect of COVID-19 vaccination on average RHR overnight in case of Moderna first dose, Pfizer-BioNTech first dose, Moderna second dose, Pfizer-BioNTech second dose respectively from left to right. Note that, during the vaccination window (five days prior to five days after vaccination), the maximum average RHR often happens at the next night or two nights after the vaccination, especially in the second dose (46% in Moderna and 54% in Pfizer-BioNTech). **C**, Distribution of symptoms reported for one week after the first (top) and second dose (bottom) of COVID-19 vaccines, Moderna and Pfizer-BioNTec. For the first dose, fatigue, poor sleep, and aches and pain are the most frequently reported symptoms with either vaccine. For the second dose, aches and pain, fatigue, and headache are the most reported symptoms in both Moderna and Pfizer-BioNTec vaccines.

The symptoms reported for one week post vaccination were recorded from the surveys (Fig. 7C). After the first dose, the most frequently occurring symptoms with either vaccine are fatigue, poor sleep, aches and pain. Post second dose, fatigue, headache, aches and pain top the list. There are some striking differences as well between Pfizer-BioNtech and Moderna. Whereas in the case of Pfizer-BioNtech, fever is reported after either dose, for Moderna, fever was not reported after the first dose but almost 60% of participants reported a fever after the second dose. Other differences can be observed from Fig 7C. Overall, these results demonstrate the presence of symptoms associated with vaccination, particularly the second dose, and that the effects of vaccination are detected using a smartwatch.

## Discussion

In this paper we introduce the first prospective real-time physiological stress detection and alerting system that can detect early-onset illness using a smartwatch. It detects COVID-19 at or prior to symptoms in 77% of the symptomatic cases and even identifies asymptomatic cases. Although the actual number of asymptomatic cases is hard to judge since most cases were not tested with RT-PCR, we nonetheless found 8 out of 10 asymptomatic cases had alerts near the test date (21 days before the diagnostic date). Detection results were similar for Fitbit and Apple Watch models. In this study, medical recommendations were not provided to participants, although doing so in future studies may provide increased healthcare value. Alerts were generated early enough (a median of 3 days prior to symptom onset for COVID-19 cases) to enable effective early self-isolation and testing.

Many of the alert-generating events detected in this study were not associated with COVID-19. The majority of annotated alerts may be attributed to other events such as poor sleep, stress, alcohol, intense exercise, travel, or other activities. In many of these cases the alerting events would be easy to self-contextualize (intense exercise, alcohol, travel) and the participants would be unlikely to take action. In other cases, such as COVID-19 negative diagnoses with symptomatic illness, follow up testing would be expected to be valuable.

The large number of unannotated alerts may be due to: a) failure to annotate an alert, b) asymptomatic infections, or c) other stressors. Since stress can trigger increases in RHR, a major feature of our detection algorithms, this approach can potentially be used to monitor mental health as well as physical health.

It is unclear why COVID-19 is not detected in all cases. Some of these individuals have a very unstable baseline, and for others the abnormal signals deviating from the baseline are not large enough to generate a signal. The inclusion of higher resolution data and/or other data types, such as heart rate variability, respiration rate, skin temperature, SpO2 changes, galvanic stress response, or other physiological features, is expected to improve detection performance for both the number of events and earliest detection time. Such data will likely help distinguish the COVID-19 cases from other non-COVID-19 events whether due to pathogenic infections, stressors, or other activities.

Notably, classic methods for illness detection have generally relied on resting oral or skin temperature and comparison of an individual to a population average instead of their individual baseline. Many COVID-19 infections do not appear to cause a fever. Moreover, skin temperature is often measured using inaccurate infrared devices and thus may not be the best method for infection detection. Resting heart rate and other longitudinal physiological measures may be valuable in conjunction with temperature measurements for early and specific disease detection. We have previously found that Lyme disease can be detected pre-symptomatically using a smartwatch and pulse oximeter, and here we demonstrate that *Mycoplasma* infection can be also identified (albeit not pre-symptomatically in this particular case). With continued development, wearable platforms, including those that use a variety of physiological parameters, can be used as a general method to monitor infectious disease, chronic inflammation-related flares, and other health-related signals to improve healthcare at both the personal and population levels and global monitoring for future pandemic outbreaks.

## Methods

### Study participation

3,246 adult individuals of 18 to 80 years of age were recruited for this study under protocol number 57022 approved by the Stanford University Institutional Review Board. Participants were invited by social media, news, and outreach to participants in previous studies. Participants registered using the REDCap survey system, and were then asked to install the study app called MyPHD (available for iOS and Android devices). The app transfers their wearable data (Fitbit via Fitbit secure OAuth 2.0 API, Apple Watch and Garmin via Healthkit repository, and other HealthKit and Google Fit compatible devices) with the Stanford research team and analysis cloud. Next, the NightSignal algorithm generated alerts (green and red) which were sent to the participants. Upon receiving alerts on the app, participants were asked to annotate events with surveys covering symptoms, activities, diagnosis, medications, and vaccination.

### Online NightSignal

A previous study on exploring whether personal sensor data can help with COVID-19 detection showed that the daily RHR on its own does not allow significant discrimination between COVID-19-positive and COVID-19-negative participants; however, it has been shown that sleep and activity data have a significant difference among the two groups ^8^. Besides, we observed significant performance improvement when overnight RHR approach is taken compared to choosing daily RHR, since this approach filters out short-range non-infection events such as stress or intense exercise during the day (Supplementary Fig. S6). For these reasons, we developed a novel algorithm based on the highest available resolution of RHR during sleep to achieve more accurate real-time detection of infection diseases such as COVID-19.

#### 1. Data pre-processing

The pre-processing stage provides consistency among different sources (i.e., Fitbit and Apple Watch) and handles missing data. The resolution of the retrieved distinct raw HRs and steps data from Fitbit and Apple Watch differs (see Supplementary Fig. S3). To calculate the RHR overnight for different devices, first we consider the heart rate records where steps are 0 and then aggregate the RHR values by calculating the average RHR during nighttime (i.e., from 12AM to 7AM). In Supplementary Fig. S4A, we show that for most subjects (over 80%), median of average RHR overnight is a stable and reliable baseline, since only after seven nights, it hits a baseline close to the baseline over three months. In the case of missing nights, we impute the values for only up to one night by calculating the average RHRs from the night before and immediately after the missed night.

#### 2. Real-time alerting

The NightSignal algorithm triggers the alerts based on the finite state machine (FSM) shown in Supplementary Fig. S2. A FSM is defined by a list of its states, its initial state, and the inputs (symbols) that trigger each transition and can produce output based on a given input and/or a state. The NightSignal state machine as depicted in Supplementary Fig. S2, contains six states and three outputs/colors (S_0_, S_1_, and S_2_ labeled with green alert, S_3_ and S_4_ labeled with yellow alert, and S_5_ labeled with red alert) and three symbols as follows: (a) A_*i*_<M_*i*_+3: For night *i*, the average RHR overnight is less than three bpm above the baseline (median of averages of RHR overnight for all nights up-to night *i*) -- The threshold of 3 was chosen since for all participants, the median of fluctuation of medians of average RHR overnight over three months was only three bpm (Supplementary Fig. S4B and Fig. S4C); (b) A_*i*_=M_*i*_+3: For night *i*, the average RHR overnight is equal to three bpm above the baseline; (c) A_*i*_>=M_*i*_+4: For night *i*, the average RHR overnight is greater than or equal to four bpm above the baseline. Transition starts from initial state S_0_ (green alert) and transition function takes one of the above six states and one of the above three symbols, and returns a state with corresponding label (alert).

### Online RHRAD

The current version of AnomalyDetect online model is built based on the previous offline model from our previous study ^7^. It uses RHR data and splits it into training data by taking the first 744 hours as a baseline (one month) and test data by taking the next one hour data, and uses a one hour sliding window to find anomalies in the test data in “real time”. If the anomalies occur frequently within 24 hours, it will automatically create either warning (yellow) or serious (red) alerts every 24 hours. Red alerts were set if the anomalies occurred continuously for more than five hours within each 24 hours period and yellow alerts were set if the anomalies occurred for one or continuously for less than five hours and green alerts were set if there were no anomalies.

### Online CuSum

We extended the CuSum online detection algorithm proposed in our previous work^7^ into the context of the real-time alerting system. Our previous work focused on the initial alarm in the purpose of early detection. In the setting of alarming, the trend of CuSum statistics was tracked in one hour resolution, the status of CuSum was evaluated in each 12 hours, and the alarm was reported each day.

In the same way as in Mishra et al^7^, the baseline was constructed from a 28-days sliding window in a personalized manner. If the data was missing more than 14 successive days, the CuSum alerting system restarted. The alerting system proceeded through chunks of data (56 days) and then the results were combined from different chunks. Under the threshold of 95%-quantile of CuSum statistics during the baseline, the difference of the standardized RHR residuals from the threshold was accumulated in each hour and the zero truncated positive difference was added to the stream of the CuSum statistics. When the CuSum statistics became significant at the first time compared to the statistics during the baseline (which serve as the null distribution), the initial alarm was called. After the initial alarm, the average CuSum statistics from each 12 hours was calculated and the CuSum entered to a yellow status. If the CuSum statistics kept increasing in two successive 12-hours intervals, a red status was recorded in the last 12-hours interval. If the CuSum statistics kept decreasing in two successive 12-hours intervals, the status was turned back to green. In the case of all missing values in the 12-hours interval, the status was recorded as NA. The alarms were sent at 9pm each day based on the latest recorded status.

## Data availability

De-identified raw heart rate and steps data used in this study can be downloaded from the following publicly available link: https://storage.googleapis.com/gbsc-gcp-project-ipop_public/COVID-19-Phase2/COVID-19-Phase2-Wearables.zip

All data will be made publicly available upon manuscript acceptance.

## Code availability

Code for the algorithms used in the manuscript are publicly available at:

Online NightSignal algorithm: https://github.com/StanfordBioinformatics/wearable-infection Online RHRAD algorithm: https://github.com/gireeshkbogu/AnomalyDetect/blob/master/scripts/rhrad_online_24hr_alerts_v6.py

Online CuSum algorithm: https://github.com/mwgrassgreen/Alarm

## Data Availability

De-identified raw heart rate and steps data used in this study can be downloaded from the following publicly available link: https://storage.googleapis.com/gbsc-gcp-project-ipop_public/COVID-19-Phase2/COVID-19-Phase2-Wearables.zip
All data will be made publicly available upon manuscript acceptance.

## Acknowledgements

This work was supported by NIH grants (1R01NR02010501 and 1S10OD023452-01) and gifts from the Flu Lab, as well as departmental funding from the Stanford Genetics department. This study was supported by the Amazon Web Services Diagnostic Development Initiative. The Google Cloud Platform costs were covered by Google for Education academic research and COVID-19 grant awards. We acknowledge the Stanford Genetics Bioinformatics Service Center (GBSC) for providing the gateway to the SCG cluster, Google Cloud Platform and Amazon Web Services for this research. The Stanford REDCap platform (http://redcap.stanford.edu) was developed and operated by the Stanford Medicine Research IT team. The REDCap platform services at Stanford are subsidized by a) Stanford School of Medicine Research Office, and b) the National Center for Research Resources and the National Center for Advancing Translational Sciences, National Institutes of Health, through grant UL1 TR001085.

## Author contributions

Study conception and design: MPS, AB, AA

Project supervision: MPS, AB

IRB Review, Participant recruitment & coordination: AC, EH, ODR, AC, AWB, JWL, BR, MPS

E-consent system (REDCap) and participant guidance: AWB, PK, AC

Wearables and survey data collection & processing: AA, AWB, ESR, AB

Software engineering (MyPHD App): AA, AB, QW, KC, RB, SP

Algorithm development and data analysis: AA, AB, GKB, MW, ESR

Cloud-based real-time alerting system: AA, AB, AAA, DC

Manuscript preparation: AA, GKB, MW, ESR, AWB, XL, AB, MPS

Manuscript review and editing: All co-authors

Funding: MPS, BR, AB, AA

## Competing interests

MPS is cofounder and a member of the scientific advisory board of Personalis, Qbio, January, SensOmics, Protos, Mirvie, Oralome. He is on the scientific advisory board of Danaher, Genapsys, and Jupiter.

## Supplementary figure and table legends

**Table1. Cohort**. Demographics, health characteristics, and COVID-19 test and vaccination info of the Cohort.

**Figure S1.**
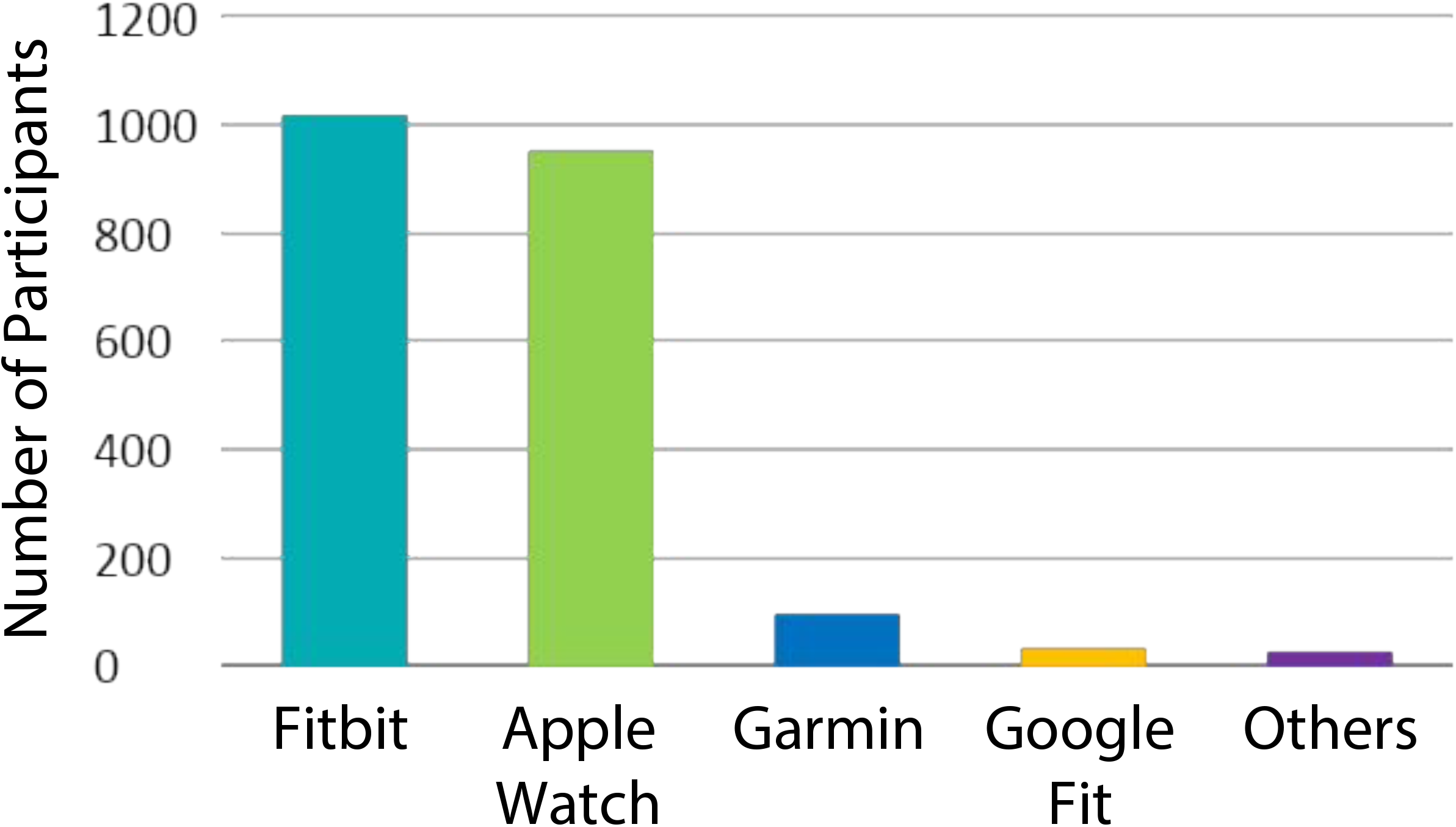
Wearable devices distribution. 2,112 of participants had a smartwatch: 1,016 wore Fitbits, 950 wore Apple Watches, 93 wore Garmin, and 53 had other devices. Note that we consider the device with the most amount of data as the main device in case of having more than one device.

**Figure S2.**
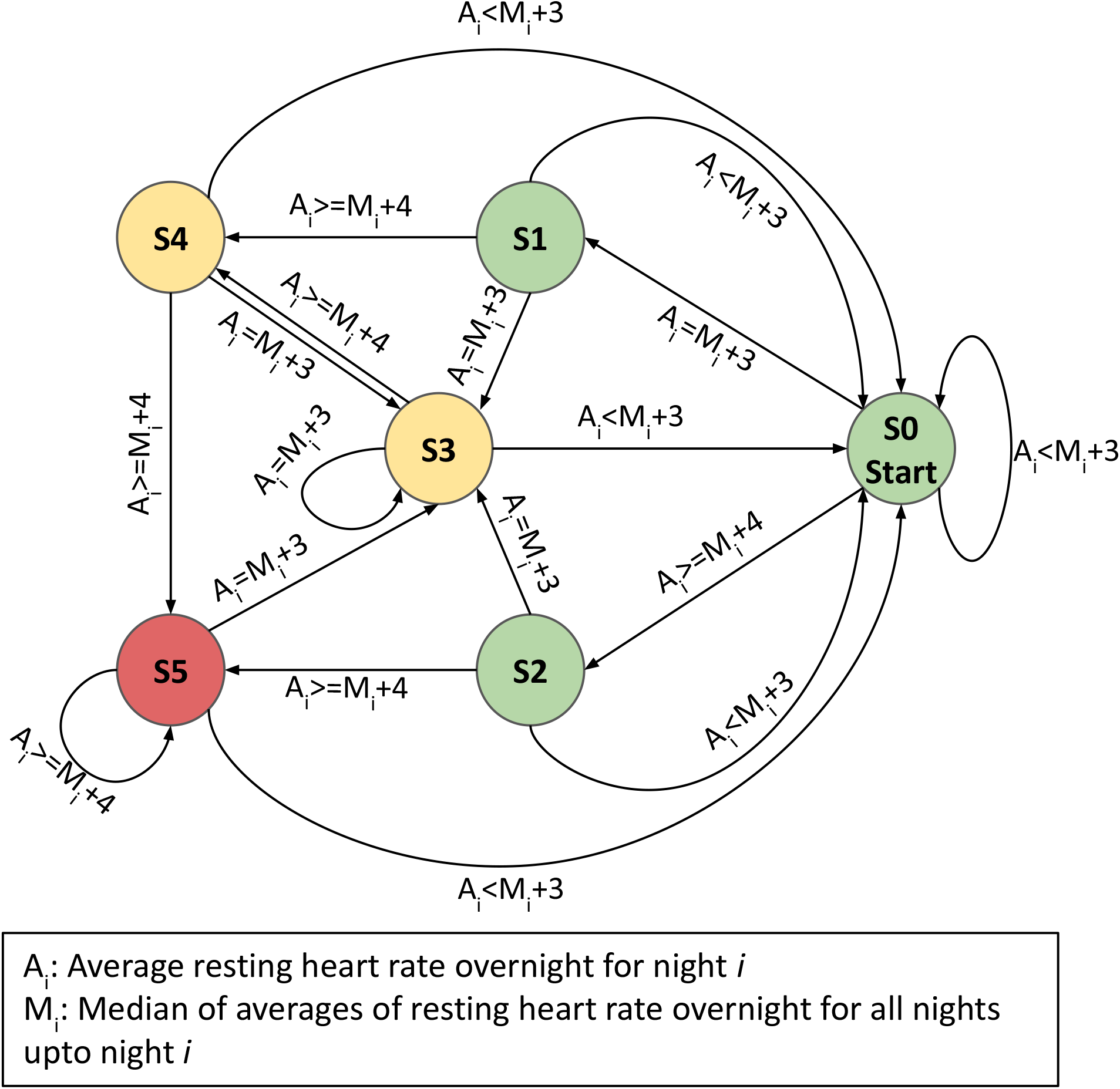
Deterministic Finite State Machine in NightSignal algorithm. The state machine consists of six states, each labeled with an alert color and three symbols for transition between states based on the current average RHR overnight and the deviation level from the baseline (streaming median of averages of RHR overnight). For example, a red alert gets triggered if for two consecutive nights, the average RHR overnight is at least four bpm above the calculated baseline.

**Figure S3.**
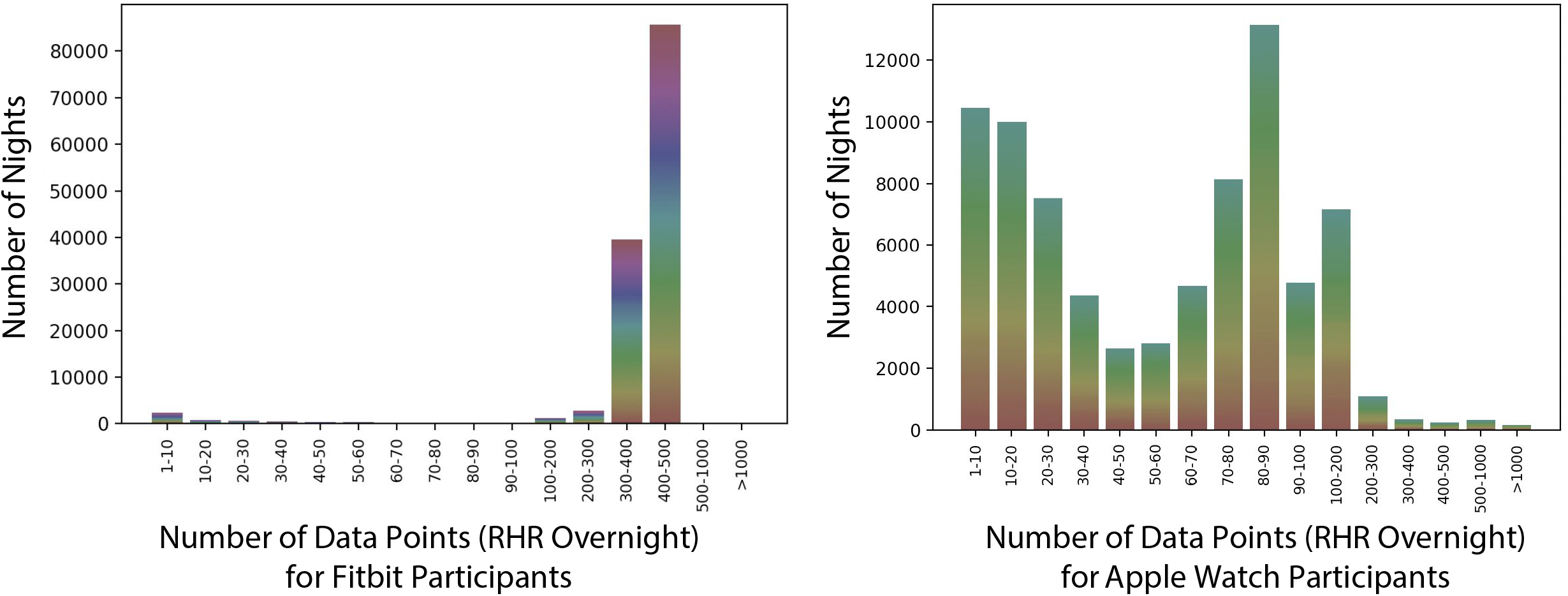
Comparing distribution of RHR overnight data in Fitbit vs. Apple Watch. To show the distribution of RHR data in Fitbit and Apple Watch, RHR overnight data points have been divided into 16 ranges (1-10, 10-20, etc) and each bar depicts the total number of nights that falls into each group. Each participant is presented with a color. Note that for the majority of participants, for most of the nights, there are 300 to 500 RHR overnight data points (i.e., almost minute resolution) in the case of Fitbit. However the range differs considerably in Apple Watch; the reason is that Apple Watch takes heart rate and step counts readings with different resolutions based on the activities. Note that in the NightSignal algorithm, we do not set a threshold for the minimum amount of data points required to aggregate per night. The first reason is that in most cases, even a very few data points are sufficient to get a proper average RHR overnight since we only consider HR records where the corresponding time interval (e.g. few minutes) for step count is zero. The second reason for that is if we do so (e.g., set the threshold to 40 data points), we will miss a significant amount of nights (e.g., first four bars in Fig. S3).

**Figure S4.**
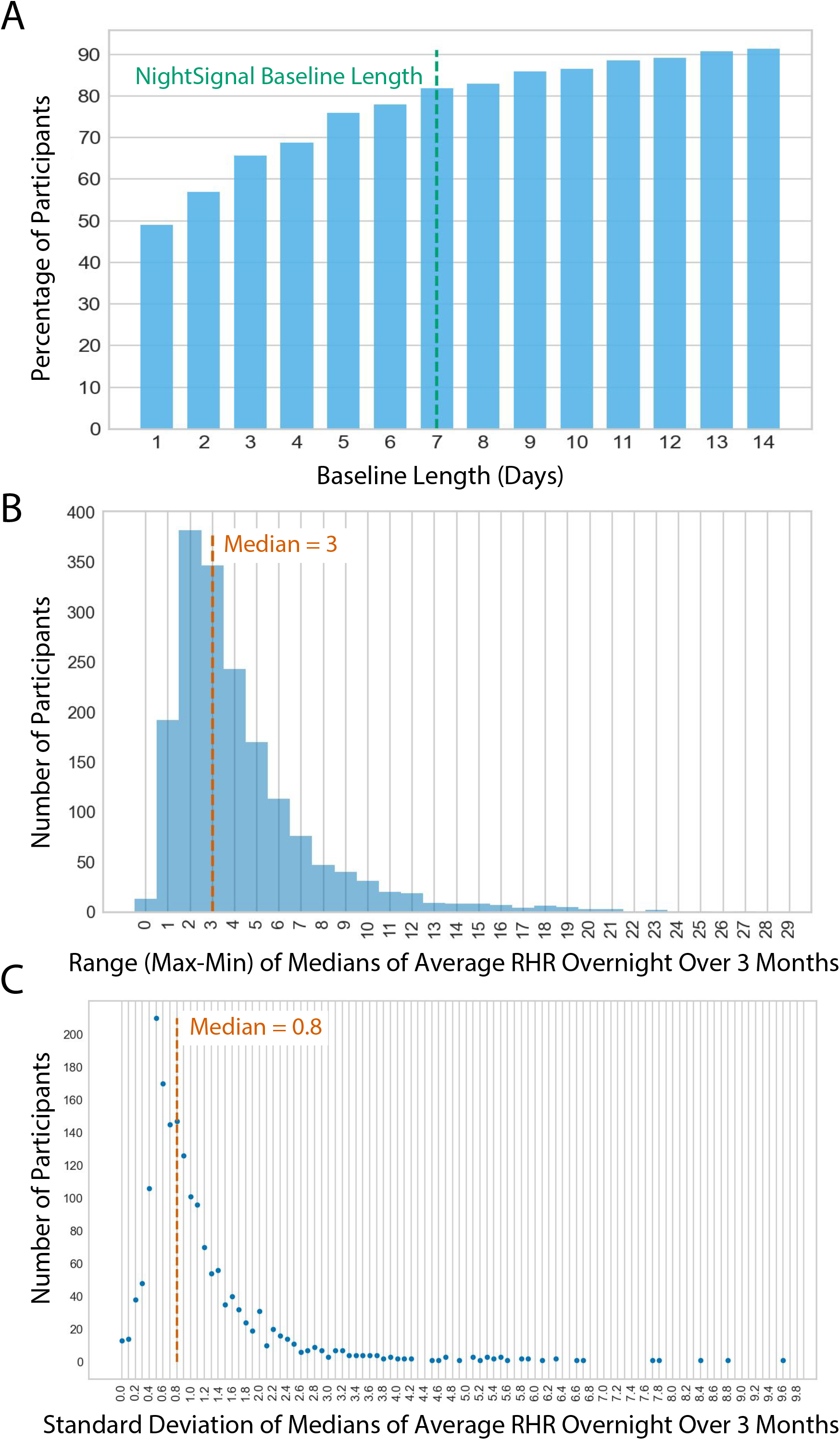
Thresholds and parameters in the NightSignal algorithm. As we discussed previously, the NightSignal algorithm uses the streaming median of average RHR overnight as the baseline. We believe that it is a proper individual healthy baseline as we show that the fluctuation is insignificant and it usually deviates due to a long-term abnormal event (e.g., infection, medication consumption, vaccination). Fig. S4A shows the minimum number of nights required to hit a baseline close to the baseline over three months (within ± two bpm from the baseline - the reason behind choosing the threshold of two is that the baseline would still remain in the green zone) for the majority of participants. As depicted in the figure, for over 80% of participants, the proper baseline was observed after only seven nights. Fig. S4B depicts the range (max-min) of medians of average RHR overnight during three months of data with the median value of only three bpm. Similarly, Quer et al.^22^ studied the variability in individual resting heart rate and showed that most subjects had a median weekly fluctuation in RHR of only three bpm. Given the fact that we only consider RHR overnight, the median fluctuation in RHR overnight is still three bpm even for a duration of three months. Similarly, Fig. S4C shows the corresponding standard deviation that is very low (0.8).

**Figure S5.**
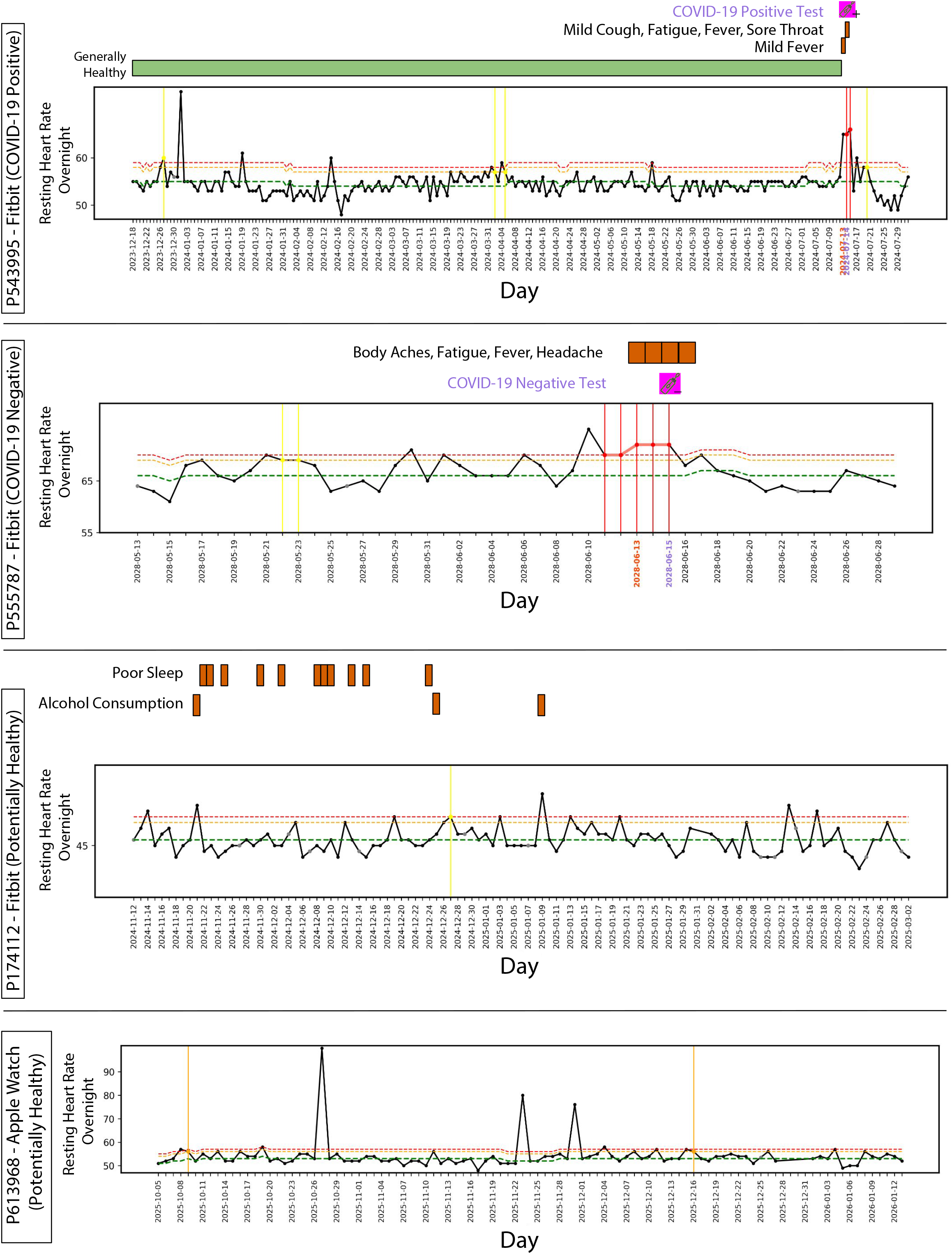
More examples of alerts for COVID-19 positive, COVID-19 negative, and potentially healthy participants. Shown are (from top to bottom): Signals for a COVID-19 positive case with mild and short-lasting symptoms. Note that this participant received zero red alerts during a healthy period of seven months and only received two red alerts starting the following night of symptoms onset date (the only period where the RHR overnight is much higher than baseline for three consecutive nights); an example for a sick participant with moderate symptoms followed by a COVID-19 negative test. Note that RHR overnight period began to increase three nights before the symptoms developed; the last two plots are the examples of healthy participants who reported no illness or symptoms of any kind during the study (except some poor sleep points for P174112). Both participants received no red alerts for over a three months healthy period (Note that for P174112, alcohol consumption affects the RHR overnight but the impact is either only for one night or not severe enough to trigger a red alert).

**Figure S6.**
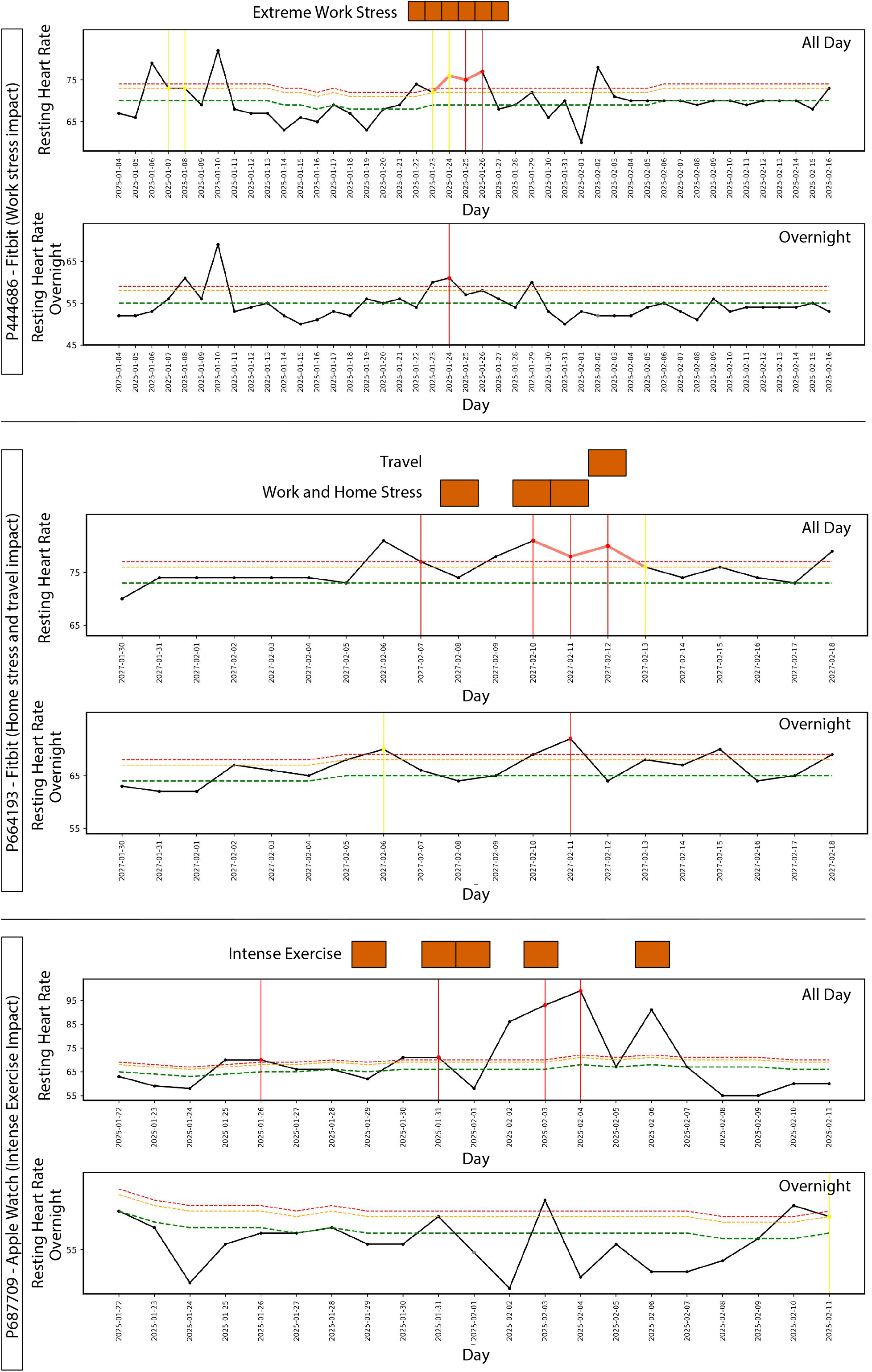
Impact of non-infectious events on NightSignal alerts (all day vs. overnight). Examples of the impact of different events (e.g., home and work stress, travel, and intense exercise) on the alerts based on two configurations: all day vs. overnight. For each participant, comparing the plots shows that the NightSignal algorithm reduces possible false positives due to non-infectious events by analyzing RHR overnight.

**Figure S7.**
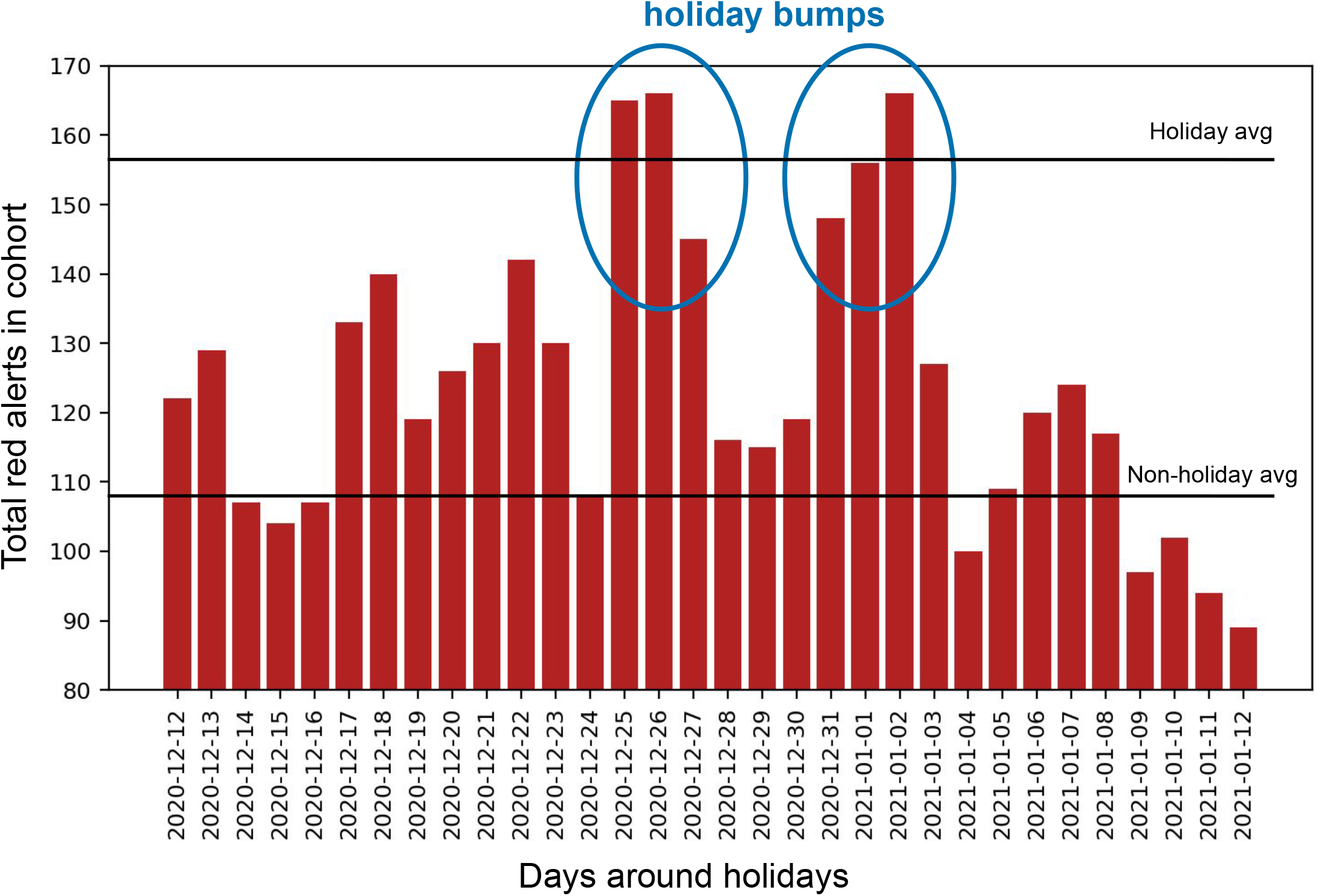
Impact of winter holidays on NightSignal alerts. As shown previously, there is a noticeable increase in the number of alerts during winter holidays -- particularly late December and beginning of January -- (“holiday bump”) due to the higher rate of travel, alcohol, entertainment, stress, and illness compared to other times of the year.

## Notes

### Author Declarations

3,246 adult individuals of 18 to 80 years of age were recruited for this study under protocol number 57022 approved by the Stanford University Institutional Review Board. Participants were invited by social media, news, and outreach to participants in previous studies. Participants registered using the REDCap survey system, and were then asked to install the study app called MyPHD (available for iOS and Android devices).

